# Global Longitudinal Active Strain Energy Density (GLASED): A Powerful Prognostic Marker in a Community-Based Cohort

**DOI:** 10.1101/2023.06.14.23291342

**Authors:** Nay Aung, David H. MacIver, Henggui Zhang, Sucharitha Chadalavada, Steffen E. Petersen

## Abstract

**BACKGROUND:** Identifying the imaging methods that best predict heart failure risk, cardiovascular adverse events and death is crucial for tailoring optimal management. Potential prognostic markers include myocardial mass, left ventricular ejection fraction, myocardial strain, stroke work, contraction fraction, pressure-strain product and a new measurement called global active longitudinal strain density (GLASED).

**OBJECTIVES:** This study sought to assess the utility of a range of potential prognostic markers of left ventricular structure and contractile function in a community-based cohort.

**METHODS:** The impact of cardiovascular magnetic resonance image-derived markers, extracted by machine learning algorithms were compared to the future risk of adverse events in a group of 44,957 UK Biobank participants.

**RESULTS:** Most markers, including the left ventricular ejection fraction, had limited prognostic value. GLASED was significantly associated with heart failure, all-cause mortality and major adverse cardiovascular events with hazard ratios of approximately 1.4.

**CONCLUSIONS:** GLASED predicted major cardiovascular adverse events and mortality with the highest hazard ratios compared with conventional markers. The routine use of GLASED is recommended for assessing prognosis.

**Graphical abstract:** 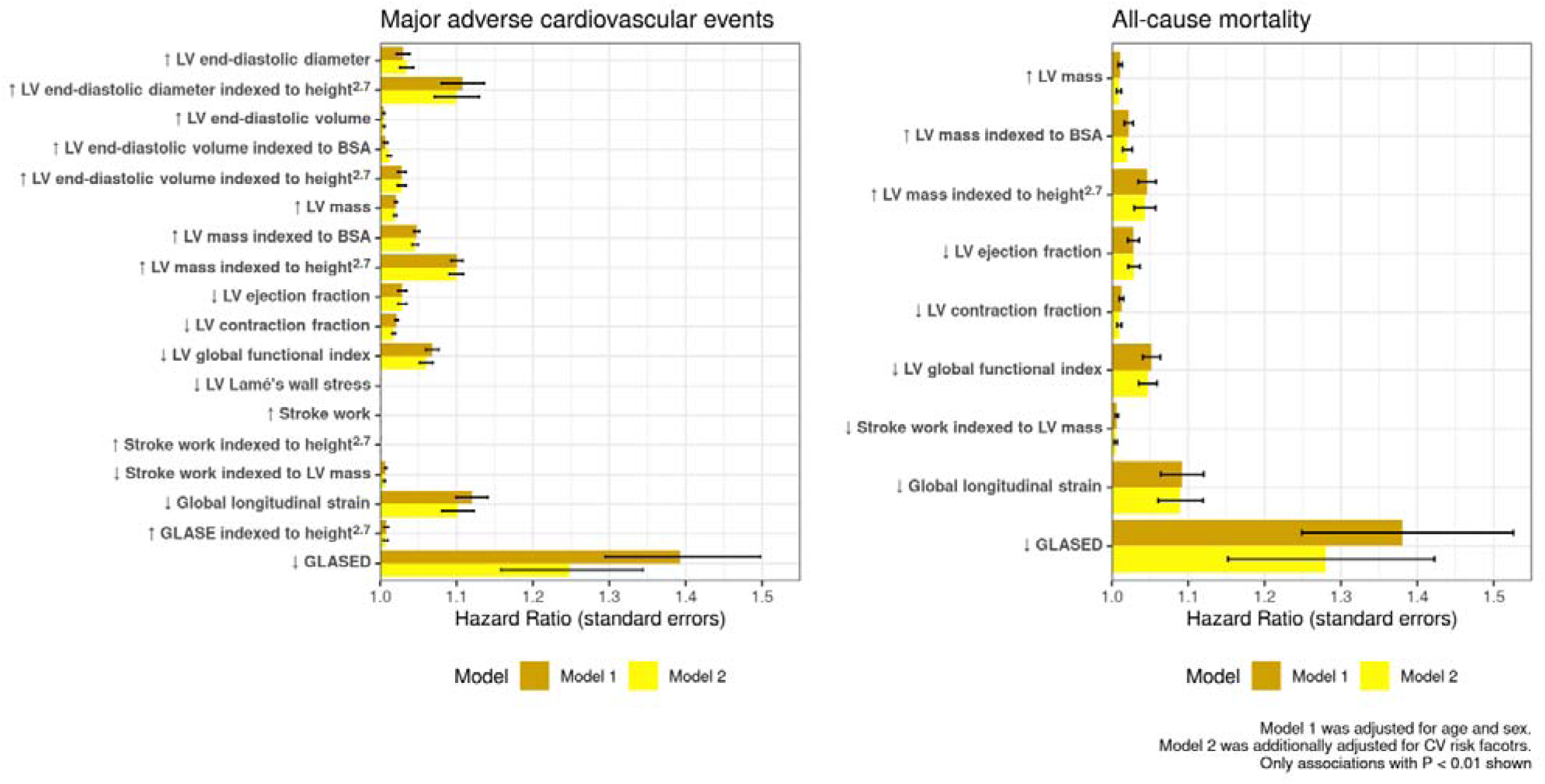

## Introduction

Cardiovascular diseases such as heart failure (HF) are a major public health issue, and identifying optimal predictive imaging markers is critical to pathophysiological understanding myocardial diseases, informing consensus documents and guiding management decisions.(1) However, the most reliable measures of left ventricular (LV) structure and function in determining the future risk of major cardiovascular adverse events (MACE), developing HF, and death are unclear.

In this study, we aimed to compare the predictive performance of the most used measures of LV structure and function, with a particular focus on a new method called global active longitudinal strain energy density (GLASED).(2)

At least 23 LV imaging measures have been advocated for the assessment of contractile function and/or risk, including left ventricular ejection fraction (LVEF),(3) end-diastolic volume,(4) LV mass,(5,6) myocardial strains,(7–9) strain rate,(10) pressure-strain product,(11) stroke work, stroke work indexed to LV mass,(2) global function index,(12) contraction fraction(13) and GLASED.(2) Although the LVEF has been widely used for risk assessment, it has serious flaws in predicting prognosis in HF syndromes.(3,14–16)

Myocardial mass associated with a greater wall thickness has been associated with a higher mortality.(5,6) Other proposed measures of contractile function, such as pressure-strain product, stroke work, stroke work indexed to LV mass, global function index, contraction fraction, have little or no outcome data.

While myocardial strain has some prognostic value,(9,10,17–19) it is constrained by its inability to consider afterload.(7,8) A higher afterload results in reduced myocardial shortening.(20) Therefore, afterload not only impacts strain interpretation it also indirectly affects LVEF since LVEF itself is influenced by strain.(21) Contractance is a new measure of contractile function derived from the area under a stress-strain curve(20) and can be estimated using GLASED(2). GLASED was introduced to overcome the limitations of strain by taking account of the effect of afterload and remodelling and estimates the mechanical energy (work done) per unit volume of myocardium during contraction. Blood pressure, wall thickness, chamber dimensions (determinants of wall stress) and myocardial strain are required for its calculation. GLASED confers a robust theoretical edge over other approaches for evaluating left ventricular systolic function because strain energy density has a strong background in engineering science. Furthermore, a recent study, in a cohort of patients referred for CMR at a regional cardiac centre, has shown GLASED to be a better predictor of expected prognosis and BNP when compared with LVEF, stroke work, stroke work per LV mass, pressure-strain product, contraction fraction, and strain.(2)

Given the theoretical benefits of GLASED, our principle apriori hypothesis was that GLASED would be a superior predictor of outcome compared with more conventional markers. We, therefore, aimed to evaluate the impact of various measures of LV structure and contractile function on mortality and morbidity in a community-based longitudinal cohort study using the UK Biobank database.

## Methods

### Study cohort

The UK Biobank is a very large prospectively recruited population study of more than 500,000 volunteers living in the United Kingdom. Detailed study protocol has been published previously.(22) The UK Biobank provides highly enriched information on demographics, lifestyle, and medical background as well as systematically collected data on physical measurements and biological samples including genomics and proteomics. An imaging enhancement sub-study with whole-body magnetic resonance was commenced in 2015 with a target sample of up to 100,000 UK Biobank participants. An overall ethical approval for UK Biobank studies was obtained from the NHS National Research Ethics Service on 17th June 2011 (Ref 11/NW/0382); this was extended on 18 June 2021 (Ref 21/NW/0157).

### Imaging analysis

In-depth information on the UK Biobank cardiovascular magnetic resonance (CMR) imaging protocol is available elsewhere.(23) In total, CMR examinations from 44,957 individuals were accessible at the time of this study. Segmentation and derivation of other LV markers were performed using a fully convolutional neural network trained on expert-annotated data in the first ≈5000 CMR studies as previously described(24,25). We excluded individuals with inadequate quality imaging data detailed in prior publications.(26,27)

### Left ventricular assessment

Global longitudinal myocardial strain (GLS) was measured using feature-tracking algorithm implemented in the CVI42 software (Prototype v5.13.7). The LV mass was calculated from myocardial volume multiplied by the density of myocardial tissue (1.05 g/ml). LV mass was indexed to body surface area (BSA) and height^2.7^.

Left ventricular global function index (LVGFI) was calculated from the following equation:(12)

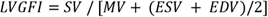

Where SV is stroke volume, MV is LV myocardial volume, ESV is end-systolic volume and EDV is end-diastolic volume.

The LV contraction fraction (LVCF) was calculated as follows(13):

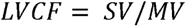

Stroke work was estimated from the product of stroke volume and systolic blood pressure.(2) Stroke work was indexed to BSA and height^2.7^.

Pressure-strain product (%mmHg) was calculated as the product of absolute strain (%) and systolic BP (mmHg).(11)

Nominal longitudinal stress (σ_l_) was calculated using the Lamé equation, as this is more accurate than Laplace’s method for thick-walled chambers, as follows:(2)

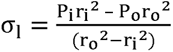

where P_i_ is inner pressure (in Pa) and equal to peak systolic pressure using a brachial cuff. P_o_ is outer (pericardial) pressure and presumed to be 0 Pa. Further, r_o_ is outer (epicardial), and r_i_ is inner (luminal or endocardial) LV radii at end-diastole, respectively.

GLASED was calculated using the following equation:(2)

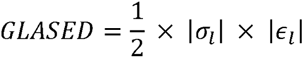

Where |*∈_l_*| is the absolute value (magnitude) of the peak GLS derived by tissue tracking to give a positive value for GLASED.

GLASE was calculated using the following equation:(2)

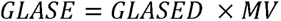

GLASE was indexed to both BSA and height^2.7^.

### Prognosis

Longitudinal follow-up of each UK Biobank participant individual was performed via linkage to Hospital Episodes Statistics (HES) data encoded in International Classification of Disease 10^th^ Revision (ICD10) classification system and national death registries. This provides information on major adverse cardiovascular events which include non-fatal or fatal myocardial infarction and stroke (MACE), incident heart failure and all-cause mortality.

### Statistical analysis

Statistical analyses were performed using R version 4.1.1.(28). We evaluated the correlations between the potential prognostic markers by calculating the Pearson correlation coefficient (r). Univariable linear regression analysis was performed to understand the relationship between LV markers and age, sex and conventional cardiovascular risk factors. For GLS, we used absolute (magnitude) values for ease of interpretation. We built Cox Proportional Hazards models to examine the associations between LV markers and heart failure, MACE or mortality. In the primary analysis, the Cox model was adjusted for age and sex (Model 1). In the secondary analyses, (i) we additionally adjusted for cardiovascular risk factors (body mass index (BMI), smoking status, regular alcohol intake, self-reported physical activity in total METs minutes per week, hypertension, diabetes mellitus and hyperlipidaemia) (Model 2) (ii) we repeated Model 1 and Model 2 in a subset of individuals with normal LVEF (>55%). We centred and scaled the variables; therefore, the effect sizes from the regression models represent per standard deviation (SD) change in the exposure variable. The effect directionality of LV markers is orientated to consistently demonstrate associations with higher risk of adverse events. A P value of less than 0.05 was considered significant.

## Results

### Demographics

The baseline characteristics of the study population are presented in Table 1. Our cohort consisted of 21,631 males and 23,326 females and an overall mean age ± standard deviation (SD) of 64 ± 8 years. The mean, SD, median, minimum and maximum values for LV markers are also shown in Table 1 and supplementary Figure S1.

**Table 1.** Demographics and main results (N=44,957)

|  | Mean (SD) or<br>n(%) | Median [min, max] |
| --- | --- | --- |
| **Age (years)** | 64.1 (7.7) | 65.0 [44.0, 82.0] |
| **Sex** |  |  |
| Male | 21631 (48.1%) |  |
| Female | 23326 (51.9%) |  |
| **Self-reported ethnicity** |  |  |
| White | 43503 (96.8%) |  |
| Asian | 484 (1.1%) |  |
| Chinese | 131 (0.3%) |  |
| Black | 293 (0.7%) |  |
| Mixed | 216 (0.5%) |  |
| Other | 235 (0.5%) |  |
| **Height (m)** | 1.7 (0.1) | 1.7 [1.3, 2.0] |
| **Weight (kg)** | 75.9 (15.1) | 74.5 [34.3, 185.0] |
| **Body mass index (kg/m<sup>2</sup>)** | 26.5 (4.4) | 25.8 [14.1, 69.6] |
| **Body surface area (m<sup>2</sup>)** | 1.9 (0.2) | 1.9 [1.2, 2.9] |
| **Systolic blood pressure (mmHg)** | 139.1 (18.7) | 137.5 [61.0, 240.5] |
| **Diastolic blood pressure (mmHg)** | 77.8 (10.5) | 77.5 [40.0, 169.0] |
| **Heart rate (bpm)** | 68.0 (11.5) | 67.0 [45.0, 156.5] |
| **Ever smoked** |  |  |
| No | 20640 (45.9%) |  |
| Yes | 23854 (53.1%) |  |
| **Physical activity (Total MET minutes per week)** | 2750.2 (2439.3) | 2039.5 [0.0, 19278.0] |
| **LV end-diastolic wall thickness (mm)** | 7.6 (1.1) | 7.5 [4.6, 18.4] |
| **LV end-diastolic diameter (mm)** | 52.0 (4.6) | 51.8 [33.8, 80.8] |
| **LV end-diastolic diameter indexed to BSA (mm/m<sup>2</sup>)** | 28.1 (2.8) | 28.1 [17.1, 45.5] |
| **LV end-diastolic diameter indexed to height<sup>2.7</sup> (mm/m<sup>2.7</sup>)** | 12.7 (1.6) | 12.6 [7.6, 23.1] |
| **LV end-diastolic volume (ml)** | 147.1 (33.8) | 142.8 [43.1, 453.1] |
| **LV end-diastolic volume indexed to BSA (ml/m<sup>2</sup>)** | 78.8 (14.1) | 77.4 [29.3, 238.8] |
| **LV end-diastolic volume indexed to height<sup>2.7</sup> (ml/m<sup>2.7</sup>)** | 35.4 (6.2) | 34.8 [15.1, 101.6] |
| **LV mass (g)** | 86.0 (22.4) | 82.7 [28.5, 287.5] |
| **LV mass indexed to BSA (g/m<sup>2</sup>)** | 45.8 (8.7) | 44.7 [17.4, 144.9] |
| **LV mass indexed to height<sup>2.7</sup> (g/m<sup>2.7</sup>)** | 20.6 (4.0) | 20.1 [8.0, 62.6] |
| **LV ejection fraction (%)** | 59.5 (6.2) | 59.7 [11.7, 82.4] |
| **LV myocardial contraction fraction** | 109.0 (20.0) | 108.2 [19.7, 277.0] |
| **LV global functional index** | 33.9 (4.5) | 33.9 [6.1, 51.9] |
| **LV Lamé's wall stress (kPa)** | 28078.7 (4578.0) | 27835.6 [8858.6, 53223.4] |
| **Pressure-strain product (mmHg%)** | 2511.8 (415.8) | 2483.2 [1066.0, 4633.4] |
| **Stroke work (mmHgml)** | 12133.8 (3308.1) | 11723.7 [2006.0, 40981.3] |
| **Stroke work indexed to BSA (mmHgml/m<sup>2</sup>)** | 6498.5 (1519.1) | 6333.9 [987.6, 18908.3] |
| **Stroke work indexed to height<sup>2.7</sup> (mmHgml/m<sup>2.7</sup>)** | 2925.6 (710.8) | 2846.2 [420.3, 9221.3] |
| **Stroke work indexed to LV mass (mmHgml/g)** | 143.4 (28.7) | 141.2 [25.2, 345.7] |
| **Global longitudinal strain (GLS) (-%)** | 18.1 (2.0) | 18.1 [12.4, 23.7] |
| **GLASE (kJ)** | 204.8 (57.3) | 197.6 [47.8, 733.2] |
| **GLASE indexed to BSA (kJ/m<sup>2</sup>)** | 109.9 (26.9) | 107.1 [32.5, 336.9] |
| **GLASE indexed to height<sup>2.7</sup> (kJ/m<sup>2.7</sup>)** | 49.4 (12.5) | 48.1 [15.5, 161.8] |
| **GLASED (kJ/m<sup>3</sup>)** | 2.6 (0.6) | 2.5 [0.8, 5.8] |

**Abbreviations:**
BMI, body mass index
BSA, body surface area
GLASE, global longitudinal active strain energy
GLASED, global longitudinal active strain energy density

### Correlation structure of LV markers

As anticipated, there was substantial correlation between different LV markers. Among LV functional markers, the strongest correlation was observed between LV ejection fraction and LVGFI (r = 0.93) Figure 1. GLASED had a high correlation with stroke work indexed to LV mass (r = 0.8) and moderate correlations with GLS, LVCF and LVGFI (r = 0.69, 0.64 and 0.5, respectively) and a low correlation with LVEF (r = 0.3). Stress and strain were not closely correlated (r=0.24).

**Figure 1.**
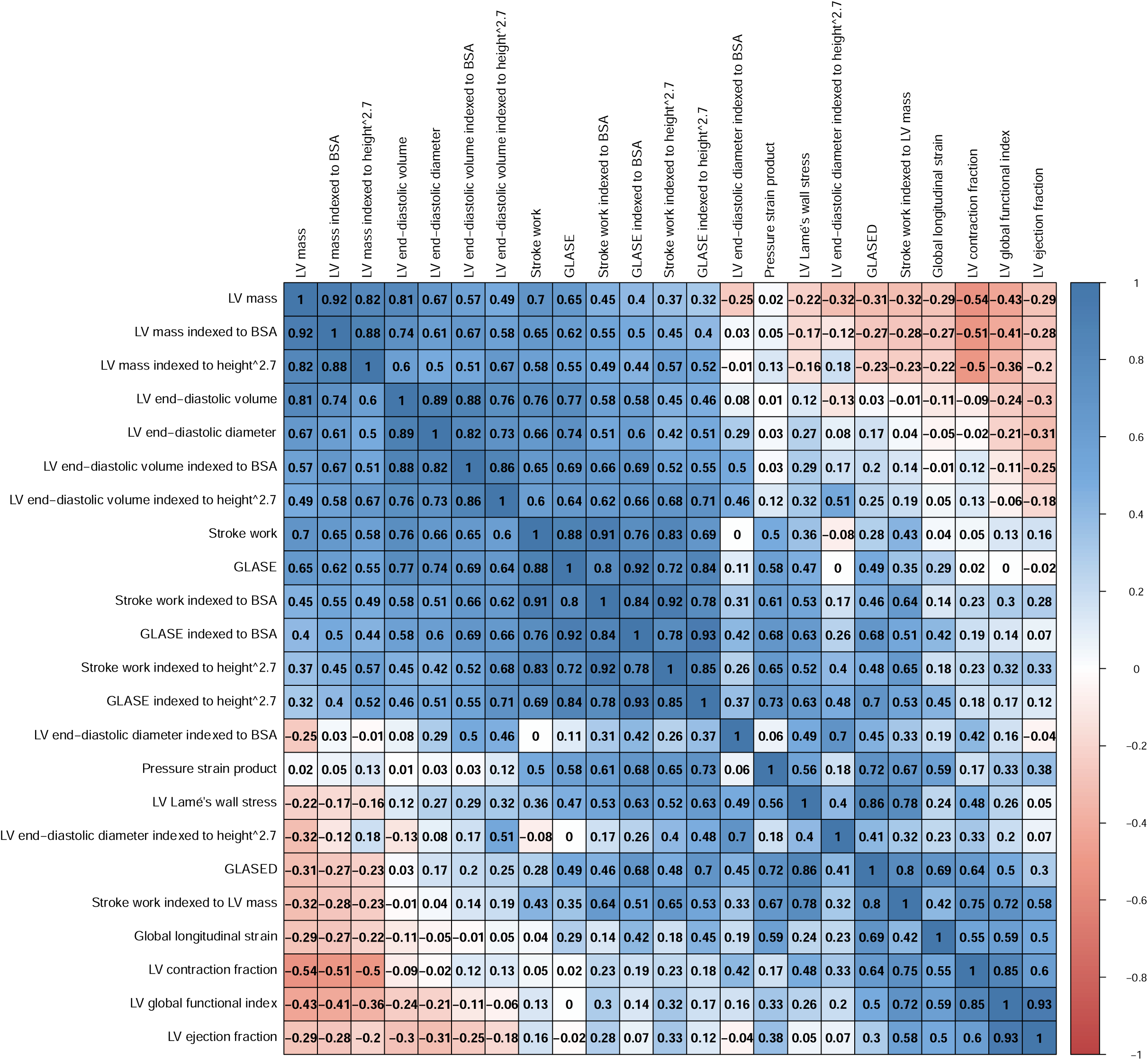
Correlation matrix of potential prognostic markers (*r* values shown)

### Relationship between LV markers and age, sex and risk factors

Age was associated with lower LV end-diastolic diameter and volume, lower LVCF, LVGFI, GLS and GLASED. With older age, higher wall stress, pressure-strain product and stroke work were observed. LV mass and end-diastolic diameter and volume were lower in females. Left ventricle ejection fraction, LVGFI and LVCF, GLS, and GLASED were significantly higher in females compared to males (Figure S2, P<0.0001). Presence of cardiovascular risk factors was consistently associated with poorer LV functional markers such as LVEF, LVCF, LVGFI, GLS and GLASED. A higher physical activity score was associated with lower LVEF but higher LVCF, GLS and GLASED (Figure S2).

### Prognostic associations with adverse outcomes

Higher LV end-diastolic diameter and volume, higher un-indexed and indexed LV mass and lower LVEF, LV contraction fraction, LVGFI, stroke work indexed to LV mass, GLS and GLASED were all associated with a higher risk of incident HF in a Cox model adjusted for age and sex (Model 1). LV end-diastolic diameter indexed to height^2.7^ had the largest effect size (hazard ratio [HR] = 1.45, 95% confidence interval [CI]: 1.33 to 1.57) followed by GLASED (HR = 1.41, 95% CI: 1.06 to 1.88) and GLS (HR = 1.30, 95% CI: 1.21 to 1.40) for every SD change. Additional adjustment with conventional cardiovascular risk factors (Model 2) rendered the heart failure risk with GLASED non-significant. Other associated LV markers from Model 1 had slightly attenuated effect sizes while retaining statistical significance in Model 2. These findings are demonstrated in Figure 2 and Supplementary Table S1.

**Figure 2.**
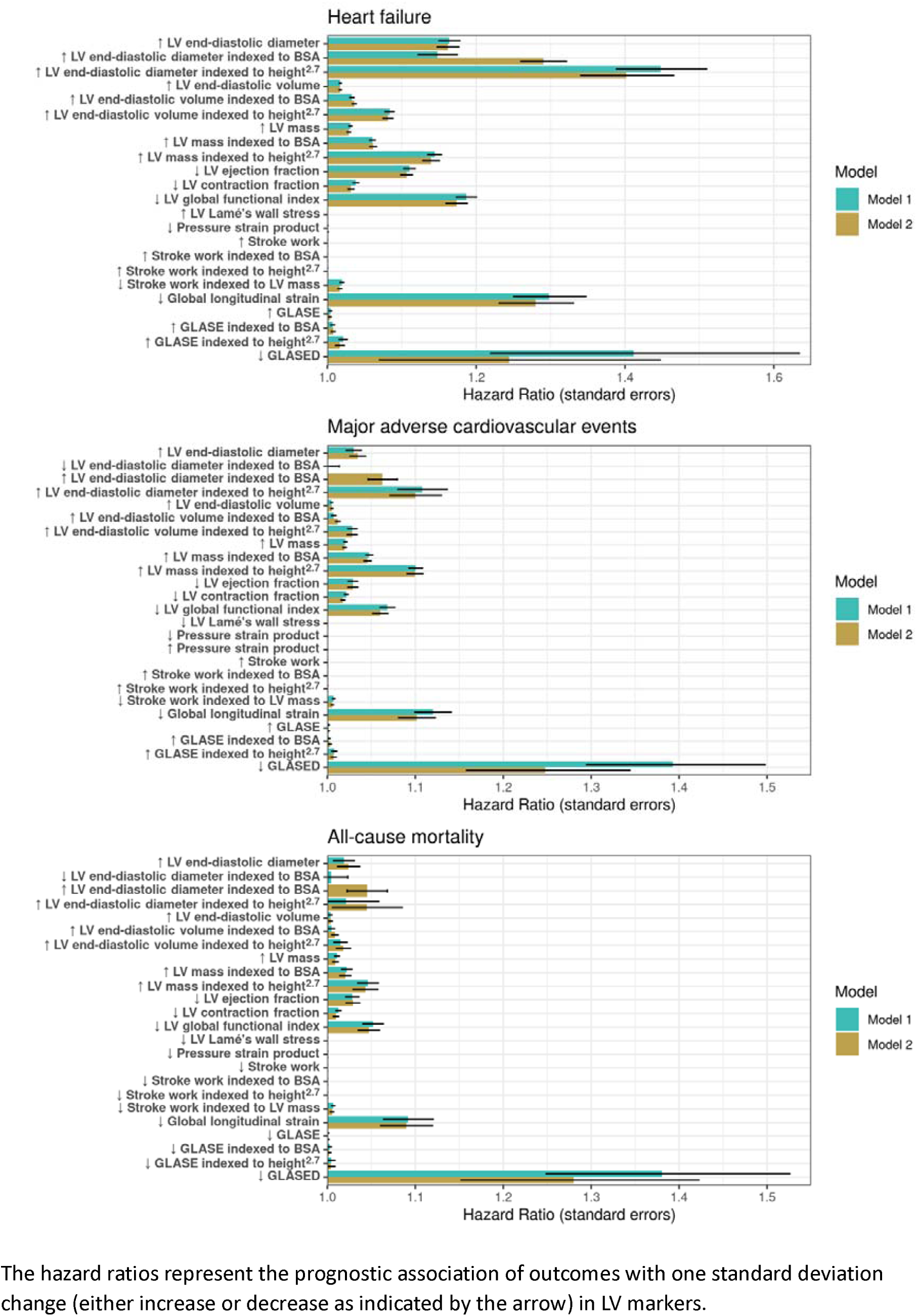
Cox proportional hazard ratios corrected for age and sex for all markers.

Higher LV mass, lower LVEF, LV contraction fraction, LVGFI, GLS and GLASED were consistently associated with a higher risk of incident MACE across both Model 1 and Model 2 (Figure S2). For every SD change in measurement, GLASED had the largest HR (1.39, CI: 1.21 to 1.61) followed by GLS (HR = 1.12, 95% CI: 1.08 to 1.16).

For all-cause mortality, higher LV mass, lower LVEF, LV contraction fraction, LVGFI, GLS and GLASED were associated with higher incident events across both Models (Figure S3). GLASED provided the largest magnitude of effect size (HR 1.38, CI: 1.13 to 1.68) in predicting death in comparison to modest effect sizes with other LV markers (mean HR ranged from 1.01 to 1.09 per SD change) as illustrated in Figure 2 and Supplementary Tables S2-S3. LV cavity size, wall stress, pressure-strain product, stroke work, stroke work indexed to BSA, stroke work indexed to height^2.7^, GLASE, GLASE indexed to BSA and GLASE indexed to height^2.7^ did not predict mortality.

In a further analysis restricted to individuals with normal LVEF (defined as >55%), GLASED remained a strong predictor of MACE and all-cause mortality with a greater HR than GLS (Model 1 HR = 1.39, CI: 1.18 to 1.64, P < 0.0001 vs HR = 1.13, CI: 1.08 to 1.18, P < 0.0001 for MACE and Model 1 HR = 1.38, CI: 1.11 to 1.73, P = 0.005 vs HR = 1.10, CI: 1.03 to 1.17, P = 0.003 for all-cause mortality, respectively), while LVEF was not significantly associated with these outcomes. Other LV markers were either not predictive or were predictive with small effect sizes (mean HR range: 1.01 to 1.09) for MACE and all-cause mortality. For HF, LVEF and GLASED were not associated with incident events, but LV mass, LV end-diastolic volume and GLS remained significantly associated in the sub-group analysis of individuals with normal LVEF. These findings are presented in Supplementary Tables S4-S6.

## Discussion

To our knowledge, this is the first study to perform a comparative and comprehensive analysis of potentially important prognostic markers of LV structure and contractile function in a large community-based cohort consisted of individuals with mostly normal LV structure (such as wall thickness and dimensions) and ejection fraction.

### Sex differences

In keeping with other studies,(29) we found a higher LVEF in females despite their lower GLS (less negative) and wall thickness. A finding that is a consequence of a reduced end-diastolic diameter.(30) LV wall stress and magnitude of GLS and GLASED were higher in females.

### Predictor markers for MACE, heart failure and all-cause mortality

Left ventricular diastolic diameter indexed to height^2.7^ had the highest HR for heart failure. An increase in LV mass only raised the risk of all outcomes slightly, although, LV mass indexed to height^2.7^ improved the prediction. LVEF predicted heart failure risk to a modest extent but was a weak marker for assessing the risk of MACE or all-cause mortality (HR = 1.03 per SD reduction). The unreliable nature of the LVEF stems from the effects of underlying variables that modulate it, such as changes in myocardial structure and strain.(21,30–33) It is recognised that a greater wall thickness or lower end-diastolic diameter increase LVEF independently of myocardial strain.(16,21,30–33) In contrast, global longitudinal strain performed better than LVEF in predicting the risk of heart failure, MACE and all-cause mortality. GLASED was the strongest marker in predicting the risk of MACE and all-cause mortality (HR ≈ 1.4). The greater HR of GLASED compared to GLS may reflect the impact of stress on the latter since GLS and stress were poorly correlated. The magnitude of effect size with GLASED for heart failure prediction was comparable to GLS (HR ratio 1.41 vs 1.30). A finding that may be a consequence of the inclination for clinicians to diagnose heart failure in the presence of dilated ventricle and/or reduced myocardial strain.

It is noteworthy that all the structural and functional markers apart from GLASED had HRs less than 1.10 for all-cause mortality. Stroke work, LV contraction fraction, global functional index and pressure-strain product were particularly disappointing as prognostic markers. A lower GLASED was associated with cardiovascular risk factors such smoking, diabetes, elevated BMI, hypertension and hyperlipidaemia because of their individual impact on wall thickness (stress) and/or strain. On the other hand, GLASED was higher with a greater level of physical activity.

Energy is often regarded as a fundamental physical quantity. GLASED is based on the principle of strain energy density which has a sound background in physics and is widely used in engineering. Apart from strain, none of the alternative techniques for assessing contractile function have an equivalent in engineering science. Information regarding strain in physics is incomplete without information about the stresses. Each of the other potential measures of contractile function, including LVEF, are indirectly derived from or a consequence of the strain energy transfer to other forms such as kinetic or pressure energy.

In summary, GLASED had the largest HR for predicting MACE and all-cause mortality, findings that are consistent with our previous study which showed that GLASED was the best predictor of expected mortality and BNP in more severe myocardial disorders i.e. hypertensive heart disease, dilated cardiomyopathy and amyloid heart disease.(2)

### Limitations

Our cohort has low pre-test probability of events given their low rates of co-morbidities and structural abnormalities. For example, there were only 2% of individuals with a LV wall thickness greater than 13 mm in the UK Biobank cohort resulting in under powering in this group. Future studies will be required to assess the risks in individuals with greater LV structural abnormalities or higher pre-test probability of cardiovascular adverse events. We would expect GLASED to be even more useful in the presence of greater structural abnormalities.

GLASED, derived using the equation quoted here, is an approximation of the myocardial longitudinal contractance derived using the area under the stress strain curve.(20) Calculating the area under the stress-strain curve requires a more sophisticated analyses that is less suitable for clinical practice.(2) However, GLASED gives comparable results to an area under curve analysis(2) and therefore was used in our study. The calculation of stress, and the derived measurements of GLASED, are prone to propagation error emphasising that accurate measurement of wall thickness and end-diastolic diameter are important. The propagation errors might have contributed towards the higher confidence intervals observed with GLASED in this study. Ambulatory blood pressure data may improve the accuracy but was not available in this cohort. No ‘diastolic function’ tests were performed as this study was principally aimed at assessing systolic function and accuracy of diastolic function assessment using CMR is questioned. Strain rates were not assessed as frame rates were too low for accurate results.

### Clinical perspective

GLS has been shown to be clinically relevant and can be measured using semi-automated techniques. GLASED could add further discriminative ability at the expense of complexity. Identifying the most powerful LV imaging marker(s) for assessing outlook is crucial to understanding pathophysiological mechanism of heart failure and exercise intolerance, guiding consensus documents and patient management as well as designing clinical intervention trials. Furthermore, the assessment of GLASED is applicable to other imaging modalities such as echocardiography.

## Conclusions

This exploratory analysis assessed the potential role of different structural and functional prognostic markers of the left ventricle and were compared with the new measure of contractile function called GLASED. We showed that a normal left ventricular ejection fraction along with multiple other measures were of limited value in predicting prognosis. Despite our cohort consisting of low-risk individuals, GLASED proved added value in the risk assessment for both major adverse cardiovascular events and mortality when compared with previously advocated methods.

## Funding section/Disclosures

Barts Charity (G-002346) contributed to fees required to access UK Biobank data [access application #2964]. SEP acknowledges the British Heart Foundation for funding the manual analysis to create a cardiovascular magnetic resonance imaging reference standard for the UK Biobank imaging resource in 5000 CMR scans (www.bhf.org.uk; PG/14/89/31194).

N.A. recognises the National Institute for Health and Care Research (NIHR) Integrated Academic Training programme which supports his Academic Clinical Lectureship post and the funding support from the Academy of Medical Sciences Clinical Lecturer Starter Grant (SGL024\1024).

SEP acknowledges support from the National Institute for Health and Care Research (NIHR) Biomedical Research Centre at Barts.

SEP and SC have received funding from the European Union’s Horizon 2020 research and innovation programme under grant agreement No 825903 (euCanSHare project).

SEP acknowledges support from the “SmartHeart” EPSRC programme grant (www.nihr.ac.uk; EP/P001009/1) and consultancy with Circle Cardiovascular Imaging Inc., Calgary, Alberta, Canada

This article is supported by the London Medical Imaging and Artificial Intelligence Centre for Value Based Healthcare (AI4VBH), which is funded from the Data to Early Diagnosis and Precision Medicine strand of the government’s Industrial Strategy Challenge Fund, managed and delivered by Innovate UK on behalf of UK Research and Innovation (UKRI). Views expressed are those of the authors and not necessarily those of the AI4VBH Consortium members, the NHS, Innovate UK, or UKRI.

The funders provided support in the form of salaries for authors as detailed above but did not have any additional role in the study design, data collection and analysis, decision to publish, or preparation of the manuscript.

## Data Availability

This research was conducted using the UK Biobank resource under access application 2964. UK Biobank will make the data available to all bona fide researchers for all types of health-related research that is in the public interest, without preferential or exclusive access for any persons. All researchers will be subject to the same application process and approval criteria as specified by UK Biobank. For more details on the access procedure, see the UK Biobank website: http://www.ukbiobank.ac.uk/register-apply/.

## Acknowledgements

This study was conducted using the UK Biobank resource under access application 2964.

We would like to thank all the participants, staff involved with planning, collection and analysis, including core lab analysis of the CMR imaging data.

## Linkage Data Acknowledgements

Please refer to the following guidance from NHS Digital and Public Health Scotland regarding the acknowledgement of data linkage.

### NHS Digital

Researchers using data from NHS Digital must cite the copyright of NHS Digital as follows: “Copyright © (year), NHS Digital. Re-used with the permission of the NHS Digital [and/or UK Biobank]. All rights reserved.” Where practicable, outputs cite the source of the data as “this work uses data provided by patients and collected by the NHS as part of their care and support.”

### Public Health Scotland

Attribution Statement - Please acknowledge National Safe Haven, providing access to the data assets and the environment, made available as part of the Data and Connectivity National Core Study Programme in papers and other research outputs. E.g. This research used data assets made available by National Safe Haven as part of the Data and Connectivity National Core Study, led by Health Data Research UK in partnership with the Office for National Statistics and funded by UK Research and Innovation (research which commenced between 1st October 2020 – 31st March 2021 grant ref MC_PC_20029; 1st April 2021 −30th September 2022 grant ref MC_PC_20058)

## Abbreviations

BMI: body mass index
BSA: body surface area
GLASE: global longitudinal active strain energy
GLASED: global longitudinal active strain energy density
GLS: global longitudinal strain
HR: hazard ratio
LV: left ventricle/ventricular
LVCF: left ventricular contraction fraction
LVEF: left ventricular ejection fraction
LVGFI: left ventricular global function index
LVMV: left ventricular muscle volume
MACE: major adverse cardiovascular events

---

**Supplementary Figure S1:**
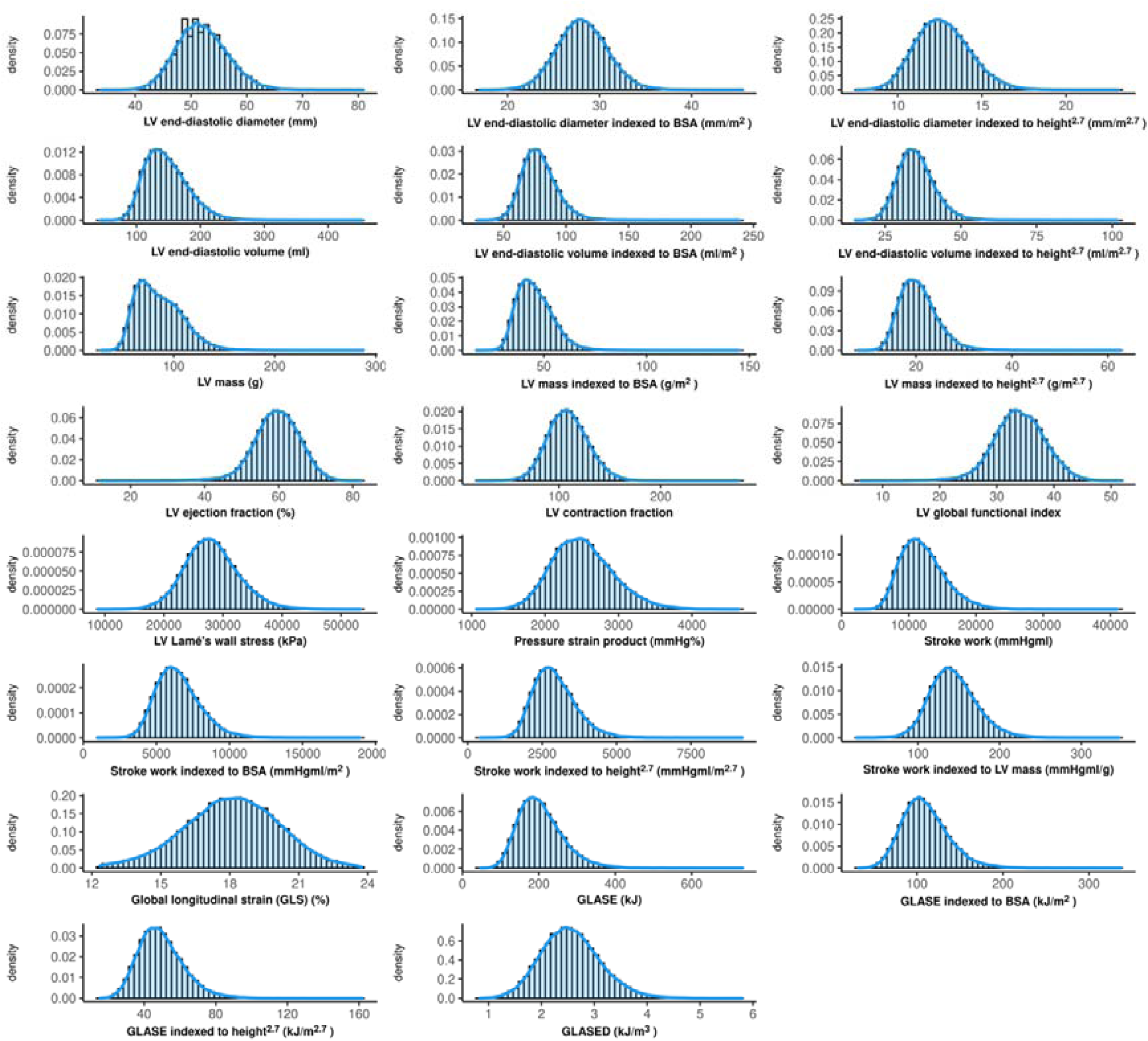
Distribution of left ventricular markers.

**Supplementary Figure S2.**
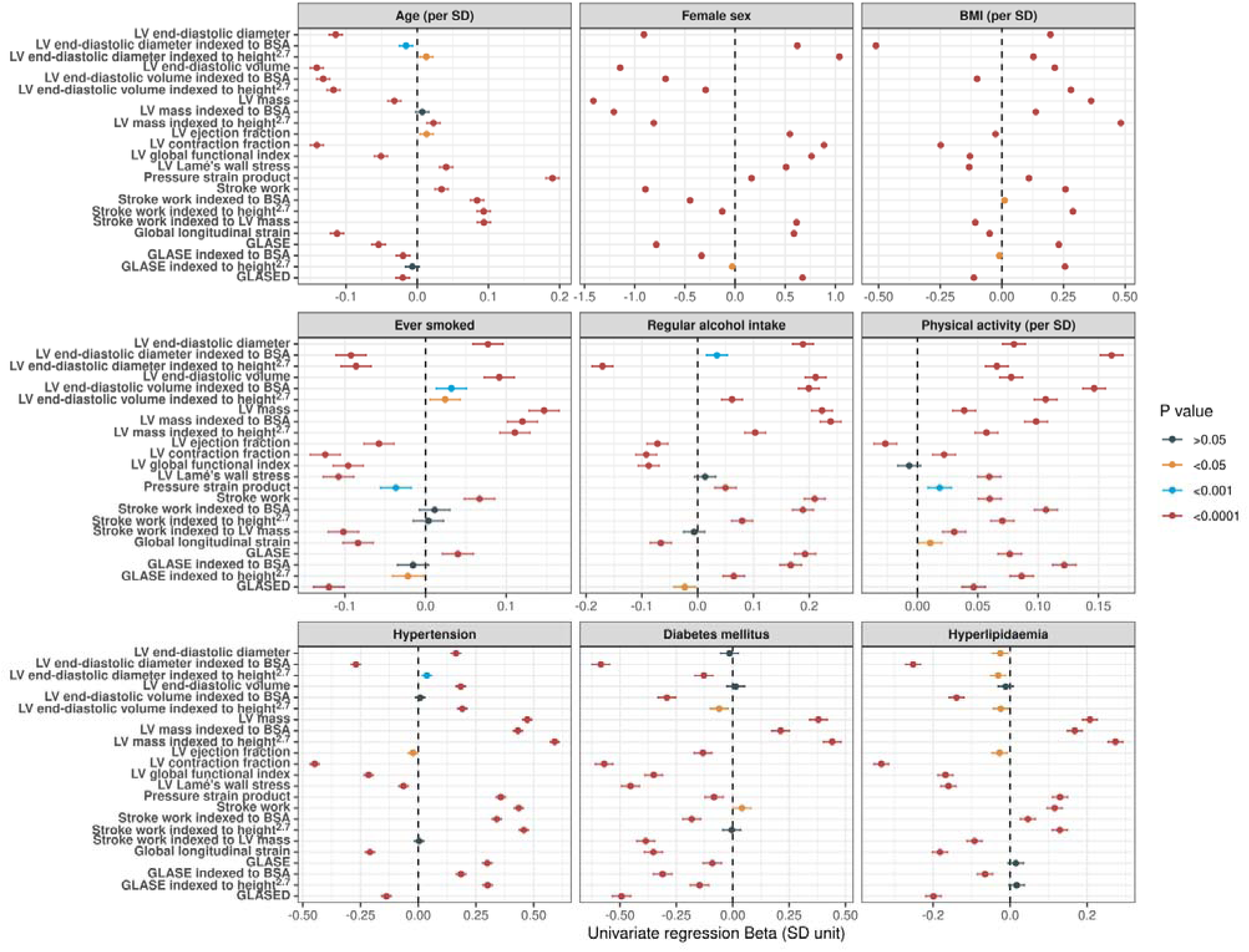
Relationship between LV markers and age, sex and risk factors (by univariate regression)

**Supplementary Table S1.** Cox regression analysis for heart failure.

| LV marker | Model 1 |  | Model 2 |  |
| --- | --- | --- | --- | --- |
|  | Hazard ratio (95% CI) | P value | Hazard ratio (95% CI) | P value |
| ↑ LV end-diastolic diameter | 1.16 (1.14 to 1.19) | <0.0001 | 1.16 (1.13 to 1.19) | <0.0001 |
| ↑ LV end-diastolic diameter indexed to BSA | 1.15 (1.10 to 1.20) | <0.0001 | 1.29 (1.23 to 1.35) | <0.0001 |
| ↑ LV end-diastolic diameter indexed to height<sup>2.7</sup> | 1.45 (1.33 to 1.57) | <0.0001 | 1.40 (1.28 to 1.53) | <0.0001 |
| ↑ LV end-diastolic volume | 1.02 (1.01 to 1.02) | <0.0001 | 1.02 (1.01 to 1.02) | <0.0001 |
| ↑ LV end-diastolic volume indexed to BSA | 1.03 (1.03 to 1.04) | <0.0001 | 1.04 (1.03 to 1.04) | <0.0001 |
| ↑ LV end-diastolic volume indexed to height<sup>2.7</sup> | 1.08 (1.07 to 1.10) | <0.0001 | 1.08 (1.07 to 1.09) | <0.0001 |
| ↑ LV mass | 1.03 (1.03 to 1.03) | <0.0001 | 1.03 (1.02 to 1.03) | <0.0001 |
| ↑ LV mass indexed to BSA | 1.06 (1.05 to 1.07) | <0.0001 | 1.06 (1.05 to 1.07) | <0.0001 |
| ↑ LV mass indexed to height<sup>2.7</sup> | 1.14 (1.13 to 1.16) | <0.0001 | 1.14 (1.12 to 1.16) | <0.0001 |
| ↓ LV ejection fraction | 1.11 (1.10 to 1.13) | <0.0001 | 1.11 (1.09 to 1.12) | <0.0001 |
| ↓ LV contraction fraction | 1.04 (1.03 to 1.05) | <0.0001 | 1.03 (1.02 to 1.04) | <0.0001 |
| ↓ LV global functional index | 1.19 (1.16 to 1.21) | <0.0001 | 1.17 (1.15 to 1.20) | <0.0001 |
| ↑ LV Lamé's wall stress | 1.00 (1.00 to 1.00) | 0.322 | 1.00 (1.00 to 1.00) | 0.026 |
| ↓ Pressure-strain product | 1.00 (1.00 to 1.00) | 0.002 | 1.00 (1.00 to 1.00) | 0.0002 |
| ↑ Stroke work | 1.00 (1.00 to 1.00) | 0.043 | 1.00 (1.00 to 1.00) | 0.392 |
| ↑ Stroke work indexed to BSA | 1.00 (1.00 to 1.00) | 0.555 | 1.00 (1.00 to 1.00) | 0.438 |
| ↑ Stroke work indexed to height<sup>2.7</sup> | 1.00 (1.00 to 1.00) | 0.017 | 1.00 (1.00 to 1.00) | 0.459 |
| ↓ Stroke work indexed to LV mass | 1.02 (1.01 to 1.02) | <0.0001 | 1.02 (1.01 to 1.02) | <0.0001 |
| ↓ Global longitudinal strain | 1.30 (1.21 to 1.40) | <0.0001 | 1.28 (1.19 to 1.38) | <0.0001 |
| ↑ GLASE | 1.00 (1.00 to 1.01) | 0.0007 | 1.00 (1.00 to 1.01) | 0.005 |
| ↑ GLASE indexed to BSA | 1.01 (1.00 to 1.01) | 0.005 | 1.01 (1.00 to 1.01) | 0.003 |
| ↑ GLASE indexed to height<sup>2.7</sup> | 1.02 (1.01 to 1.03) | <0.0001 | 1.02 (1.01 to 1.03) | 0.003 |
| ↓ GLASED | 1.41 (1.06 to 1.88) | 0.019 | 1.24 (0.93 to 1.67) | 0.147 |

**Supplementary Table S2.** Cox regression analysis for major adverse cardiovascular events.

| LV marker | Model 1 |  | Model 2 |  |
| --- | --- | --- | --- | --- |
|  | Hazard ratio (95% CI) | P value | Hazard ratio (95% CI) | P value |
| ↑ LV end-diastolic diameter | 1.03 (1.01 to 1.05) | 0.0004 | 1.03 (1.02 to 1.05) | <0.0001 |
| ↓ LV end-diastolic diameter indexed to BSA | 1.00 (0.97 to 1.03) | 0.999 | 1.06 (1.03 to 1.10) | <0.0001 |
| ↑ LV end-diastolic diameter indexed to height<sup>2,7</sup> | 1.11 (1.05 to 1.16) | <0.0001 | 1.10 (1.04 to 1.16) | 0.0003 |
| ↑ LV end-diastolic volume | 1.00 (1.00 to 1.01) | <0.0001 | 1.01 (1.00 to 1.01) | <0.0001 |
| ↑ LV end-diastolic volume indexed to BSA | 1.01 (1.00 to 1.01) | 0.003 | 1.01 (1.01 to 1.02) | <0.0001 |
| ↑ LV end-diastolic volume indexed to height<sup>2,7</sup> | 1.03 (1.02 to 1.04) | <0.0001 | 1.03 (1.02 to 1.04) | <0.0001 |
| ↑ LV mass | 1.02 (1.02 to 1.02) | <0.0001 | 1.02 (1.02 to 1.02) | <0.0001 |
| ↑ LV mass indexed to BSA | 1.05 (1.04 to 1.06) | <0.0001 | 1.05 (1.04 to 1.05) | <0.0001 |
| ↑ LV mass indexed to height<sup>2,7</sup> | 1.10 (1.09 to 1.12) | <0.0001 | 1.10 (1.08 to 1.12) | <0.0001 |
| ↓ LV ejection fraction | 1.03 (1.02 to 1.04) | <0.0001 | 1.03 (1.02 to 1.04) | <0.0001 |
| ↓ LV contraction fraction | 1.02 (1.02 to 1.03) | <0.0001 | 1.02 (1.01 to 1.02) | <0.0001 |
| ↓ LV global functional index | 1.07 (1.05 to 1.08) | <0.0001 | 1.06 (1.04 to 1.08) | <0.0001 |
| ↓ LV Lamé's wall stress | 1.00 (1.00 to 1.00) | 0.004 | 1.00 (1.00 to 1.00) | 0.259 |
| ↑ Pressure-strain product | 1.00 (1.00 to 1.00) | 0.664 | 1.00 (1.00 to 1.00) | 0.825 |
| ↑ Stroke work | 1.00 (1.00 to 1.00) | 0.0008 | 1.00 (1.00 to 1.00) | 0.007 |
| ↑ Stroke work indexed to BSA | 1.00 (1.00 to 1.00) | 0.019 | 1.00 (1.00 to 1.00) | 0.004 |
| ↑ Stroke work indexed to height<sup>2,7</sup> | 1.00 (1.00 to 1.00) | <0.0001 | 1.00 (1.00 to 1.00) | 0.003 |
| ↓ Stroke work indexed to LV mass | 1.01 (1.00 to 1.01) | <0.0001 | 1.01 (1.00 to 1.01) | <0.0001 |
| ↓ Global longitudinal strain | 1.12 (1.08 to 1.16) | <0.0001 | 1.10 (1.06 to 1.14) | <0.0001 |
| ↑ GLASE | 1.00 (1.00 to 1.00) | 0.021 | 1.00 (1.00 to 1.00) | 0.028 |
| ↑ GLASE indexed to BSA | 1.00 (1.00 to 1.00) | 0.192 | 1.00 (1.00 to 1.01) | 0.022 |
| ↑ GLASE indexed to height<sup>2,7</sup> | 1.01 (1.00 to 1.01) | 0.004 | 1.01 (1.00 to 1.01) | 0.016 |
| ↓ GLASED | 1.39 (1.21 to 1.61) | <0.0001 | 1.25 (1.08 to 1.44) | 0.003 |

**Supplementary Table S3.** Cox regression analysis for all-cause mortality.

|  | **Model 1** |  | **Model 2** |  |
| --- | --- | --- | --- | --- |
| **LV marker** | **Hazard ratio (95% CI)** | **P value** | **Hazard ratio (95% CI)** | **P value** |
| ↑ LV end-diastolic diameter | 1.02 (1.00 to 1.04) | 0.109 | 1.02 (1.00 to 1.05) | 0.051 |
| ↓ LV end-diastolic diameter indexed to BSA | 1.00 (0.97 to 1.04) | 0.818 | 1.05 (1.00 to 1.09) | 0.044 |
| ↑ LV end-diastolic diameter indexed to height<sup>2.7</sup> | 1.02 (0.95 to 1.10) | 0.552 | 1.04 (0.97 to 1.13) | 0.249 |
| ↑ LV end-diastolic volume | 1.00 (1.00 to 1.01) | 0.033 | 1.00 (1.00 to 1.01) | 0.030 |
| ↑ LV end-diastolic volume indexed to BSA | 1.00 (1.00 to 1.01) | 0.165 | 1.01 (1.00 to 1.02) | 0.016 |
| ↑ LV end-diastolic volume indexed to height<sup>2.7</sup> | 1.01 (1.00 to 1.03) | 0.044 | 1.02 (1.00 to 1.03) | 0.021 |
| ↑ LV mass | 1.01 (1.01 to 1.02) | <0.0001 | 1.01 (1.00 to 1.02) | 0.002 |
| ↑ LV mass indexed to BSA | 1.02 (1.01 to 1.03) | 0.0001 | 1.02 (1.01 to 1.03) | 0.001 |
| ↑ LV mass indexed to height<sup>2.7</sup> | 1.05 (1.02 to 1.07) | <0.0001 | 1.04 (1.02 to 1.07) | 0.002 |
| ↓ LV ejection fraction | 1.03 (1.01 to 1.04) | 0.0001 | 1.03 (1.01 to 1.04) | 0.0001 |
| ↓ LV contraction fraction | 1.01 (1.01 to 1.02) | <0.0001 | 1.01 (1.00 to 1.02) | 0.001 |
| ↓ LV global functional index | 1.05 (1.03 to 1.07) | <0.0001 | 1.05 (1.02 to 1.07) | <0.0001 |
| ↓ LV Lamé's wall stress | 1.00 (1.00 to 1.00) | 0.059 | 1.00 (1.00 to 1.00) | 0.476 |
| ↓ Pressure-strain product | 1.00 (1.00 to 1.00) | 0.118 | 1.00 (1.00 to 1.00) | 0.087 |
| ↓ Stroke work | 1.00 (1.00 to 1.00) | 0.658 | 1.00 (1.00 to 1.00) | 0.439 |
| ↓ Stroke work indexed to BSA | 1.00 (1.00 to 1.00) | 0.240 | 1.00 (1.00 to 1.00) | 0.408 |
| ↓ Stroke work indexed to height<sup>2.7</sup> | 1.00 (1.00 to 1.00) | 0.460 | 1.00 (1.00 to 1.00) | 0.361 |
| ↓ Stroke work indexed to LV mass | 1.01 (1.00 to 1.01) | 0.0002 | 1.01 (1.00 to 1.01) | 0.005 |
| ↓ Global longitudinal strain | 1.09 (1.04 to 1.15) | 0.0007 | 1.09 (1.03 to 1.15) | 0.001 |
| ↓ GLASE | 1.00 (1.00 to 1.00) | 0.480 | 1.00 (1.00 to 1.00) | 0.436 |
| ↓ GLASE indexed to BSA | 1.00 (1.00 to 1.01) | 0.169 | 1.00 (1.00 to 1.01) | 0.335 |
| ↓ GLASE indexed to height<sup>2.7</sup> | 1.00 (1.00 to 1.01) | 0.269 | 1.00 (1.00 to 1.01) | 0.352 |
| ↓ GLASED | 1.38 (1.13 to 1.68) | 0.001 | 1.28 (1.04 to 1.57) | 0.019 |

**Supplementary Table S4.** Cox regression analysis for heart failure in normal LVEF (>55%)

| LV marker | Model 1 |  | Model 2 |  |
| --- | --- | --- | --- | --- |
|  | Hazard ratio (95% CI) | P value | Hazard ratio (95% CI) | P value |
| ↑ LV end-diastolic diameter | 1.10 (1.05 to 1.15) | <0.0001 | 1.08 (1.03 to 1.14) | 0.001 |
| ↑ LV end-diastolic diameter indexed to BSA | 1.00 (0.93 to 1.08) | 0.929 | 1.18 (1.08 to 1.28) | 0.0002 |
| ↑ LV end-diastolic diameter indexed to height<sup>2,7</sup> | 1.35 (1.19 to 1.53) | <0.0001 | 1.25 (1.09 to 1.44) | 0.001 |
| ↑ LV end-diastolic volume | 1.01 (1.01 to 1.02) | <0.0001 | 1.01 (1.01 to 1.02) | 0.0004 |
| ↑ LV end-diastolic volume indexed to BSA | 1.02 (1.01 to 1.04) | 0.002 | 1.03 (1.02 to 1.04) | <0.0001 |
| ↑ LV end-diastolic volume indexed to height<sup>2,7</sup> | 1.08 (1.06 to 1.11) | <0.0001 | 1.07 (1.04 to 1.10) | <0.0001 |
| ↑ LV mass | 1.03 (1.02 to 1.04) | <0.0001 | 1.02 (1.01 to 1.03) | 0.0001 |
| ↑ LV mass indexed to BSA | 1.06 (1.04 to 1.08) | <0.0001 | 1.05 (1.03 to 1.07) | <0.0001 |
| ↑ LV mass indexed to height<sup>2,7</sup> | 1.16 (1.12 to 1.20) | <0.0001 | 1.11 (1.06 to 1.16) | <0.0001 |
| ↓ LV ejection fraction | 1.01 (0.96 to 1.05) | 0.791 | 1.01 (0.96 to 1.05) | 0.762 |
| ↓ LV contraction fraction | 1.01 (1.00 to 1.02) | 0.029 | 1.00 (0.99 to 1.01) | 0.887 |
| ↓ LV global functional index | 1.05 (0.99 to 1.11) | 0.103 | 1.00 (0.94 to 1.06) | 0.958 |
| ↑ LV Lamé's wall stress | 1.00 (1.00 to 1.00) | 0.844 | 1.00 (1.00 to 1.00) | 0.249 |
| ↓ Pressure-strain loop | 1.00 (1.00 to 1.00) | 0.989 | 1.00 (1.00 to 1.00) | 0.703 |
| ↑ Stroke work | 1.00 (1.00 to 1.00) | <0.0001 | 1.00 (1.00 to 1.00) | 0.003 |
| ↑ Stroke work indexed to BSA | 1.00 (1.00 to 1.00) | 0.002 | 1.00 (1.00 to 1.00) | 0.0006 |
| ↑ Stroke work indexed to height<sup>2,7</sup> | 1.00 (1.00 to 1.00) | <0.0001 | 1.00 (1.00 to 1.00) | 0.0005 |
| ↓ Stroke work indexed to LV mass | 1.00 (1.00 to 1.01) | 0.570 | 1.00 (0.99 to 1.01) | 0.595 |
| ↓ Global longitudinal strain | 1.14 (1.03 to 1.26) | 0.011 | 1.10 (0.99 to 1.22) | 0.085 |
| ↑ GLASE | 1.00 (1.00 to 1.01) | 0.010 | 1.00 (1.00 to 1.01) | 0.064 |
| ↑ GLASE indexed to BSA | 1.01 (1.00 to 1.01) | 0.063 | 1.01 (1.00 to 1.01) | 0.028 |
| ↑ GLASE indexed to height<sup>2,7</sup> | 1.02 (1.01 to 1.04) | 0.0006 | 1.02 (1.00 to 1.03) | 0.017 |
| ↓ GLASED | 1.17 (0.81 to 1.68) | 0.401 | 1.03 (0.71 to 1.49) | 0.888 |

**Supplementary Table S5.** Cox regression analysis for major adverse cardiovascular events in normal LVEF (>55%)

| LV marker | Model 1 |  | Model 2 |  |
| --- | --- | --- | --- | --- |
|  | Hazard ratio (95% CI) | P value | Hazard ratio (95% CI) | P value |
| ↑ LV end-diastolic diameter | 1.01 (0.99 to 1.03) | 0.465 | 1.01 (0.99 to 1.04) | 0.247 |
| ↓ LV end-diastolic diameter indexed to BSA | 1.04 (1.00 to 1.07) | 0.024 | 1.01 (0.97 to 1.05) | 0.658 |
| ↑ LV end-diastolic diameter indexed to height<sup>2,7</sup> | 1.03 (0.97 to 1.09) | 0.360 | 1.02 (0.95 to 1.08) | 0.628 |
| ↑ LV end-diastolic volume | 1.00 (1.00 to 1.00) | 0.275 | 1.00 (1.00 to 1.01) | 0.228 |
| ↓ LV end-diastolic volume indexed to BSA | 1.00 (0.99 to 1.01) | 0.731 | 1.00 (1.00 to 1.01) | 0.282 |
| ↑ LV end-diastolic volume indexed to height<sup>2,7</sup> | 1.01 (1.00 to 1.02) | 0.137 | 1.01 (0.99 to 1.02) | 0.251 |
| ↑ LV mass | 1.02 (1.01 to 1.02) | <0.0001 | 1.02 (1.01 to 1.02) | <0.0001 |
| ↑ LV mass indexed to BSA | 1.04 (1.03 to 1.05) | <0.0001 | 1.04 (1.03 to 1.05) | <0.0001 |
| ↑ LV mass indexed to height<sup>2,7</sup> | 1.09 (1.07 to 1.11) | <0.0001 | 1.09 (1.06 to 1.11) | <0.0001 |
| ↑ LV ejection fraction | 1.00 (0.98 to 1.02) | 0.843 | 1.01 (0.99 to 1.03) | 0.556 |
| ↓ LV contraction fraction | 1.02 (1.01 to 1.03) | <0.0001 | 1.02 (1.01 to 1.02) | <0.0001 |
| ↓ LV global functional index | 1.06 (1.03 to 1.09) | <0.0001 | 1.05 (1.02 to 1.08) | 0.0003 |
| ↓ LV Lamé's wall stress | 1.00 (1.00 to 1.00) | 0.012 | 1.00 (1.00 to 1.00) | 0.200 |
| ↑ Pressure-strain loop | 1.00 (1.00 to 1.00) | 0.179 | 1.00 (1.00 to 1.00) | 0.383 |
| ↑ Stroke work | 1.00 (1.00 to 1.00) | 0.0001 | 1.00 (1.00 to 1.00) | 0.002 |
| ↑ Stroke work indexed to BSA | 1.00 (1.00 to 1.00) | 0.002 | 1.00 (1.00 to 1.00) | 0.001 |
| ↑ Stroke work indexed to height<sup>2,7</sup> | 1.00 (1.00 to 1.00) | <0.0001 | 1.00 (1.00 to 1.00) | 0.001 |
| ↓ Stroke work indexed to LV mass | 1.00 (1.00 to 1.01) | 0.011 | 1.00 (1.00 to 1.01) | 0.089 |
| ↓ Global longitudinal strain | 1.13 (1.08 to 1.18) | <0.0001 | 1.11 (1.05 to 1.16) | <0.0001 |
| ↑ GLASE | 1.00 (1.00 to 1.00) | 0.116 | 1.00 (1.00 to 1.00) | 0.174 |
| ↑ GLASE indexed to BSA | 1.00 (1.00 to 1.00) | 0.510 | 1.00 (1.00 to 1.01) | 0.178 |
| ↑ GLASE indexed to height<sup>2,7</sup> | 1.01 (1.00 to 1.01) | 0.060 | 1.00 (1.00 to 1.01) | 0.160 |
| ↓ GLASED | 1.39 (1.18 to 1.64) | <0.0001 | 1.25 (1.06 to 1.48) | 0.009 |

**Supplementary Table S6.** Cox regression analysis for all-cause mortality in normal LVEF (>55%)

| LV parameter | Model 1 |  | Model 2 |  |
| --- | --- | --- | --- | --- |
|  | Hazard ratio (95% CI) | P value | Hazard ratio (95% CI) | P value |
| ↓ LV end-diastolic diameter | 1.01 (0.98 to 1.04) | 0.592 | 1.00 (0.97 to 1.03) | 0.901 |
| ↓ LV end-diastolic diameter indexed to BSA | 1.04 (1.00 to 1.09) | 0.080 | 1.01 (0.96 to 1.07) | 0.700 |
| ↓ LV end-diastolic diameter indexed to height<sup>2.7</sup> | 1.07 (0.98 to 1.17) | 0.119 | 1.05 (0.95 to 1.15) | 0.346 |
| ↓ LV end-diastolic volume | 1.00 (1.00 to 1.01) | 0.610 | 1.00 (1.00 to 1.01) | 0.697 |
| ↓ LV end-diastolic volume indexed to BSA | 1.01 (1.00 to 1.02) | 0.176 | 1.00 (0.99 to 1.01) | 0.550 |
| ↓ LV end-diastolic volume indexed to height<sup>2.7</sup> | 1.01 (0.99 to 1.03) | 0.239 | 1.01 (0.99 to 1.03) | 0.403 |
| ↑ LV mass | 1.01 (1.00 to 1.01) | 0.029 | 1.01 (1.00 to 1.01) | 0.122 |
| ↑ LV mass indexed to BSA | 1.01 (1.00 to 1.03) | 0.104 | 1.01 (1.00 to 1.03) | 0.159 |
| ↑ LV mass indexed to height<sup>2.7</sup> | 1.02 (0.99 to 1.06) | 0.124 | 1.02 (0.98 to 1.06) | 0.271 |
| ↓ LV ejection fraction | 1.01 (0.98 to 1.04) | 0.513 | 1.01 (0.98 to 1.04) | 0.642 |
| ↓ LV contraction fraction | 1.01 (1.00 to 1.02) | 0.001 | 1.01 (1.00 to 1.02) | 0.032 |
| ↓ LV global functional index | 1.04 (1.01 to 1.08) | 0.024 | 1.03 (0.99 to 1.07) | 0.128 |
| ↓ LV Lamé's wall stress | 1.00 (1.00 to 1.00) | 0.052 | 1.00 (1.00 to 1.00) | 0.258 |
| ↓ Pressure-strain loop | 1.00 (1.00 to 1.00) | 0.234 | 1.00 (1.00 to 1.00) | 0.205 |
| ↑ Stroke work | 1.00 (1.00 to 1.00) | 0.994 | 1.00 (1.00 to 1.00) | 0.990 |
| ↓ Stroke work indexed to BSA | 1.00 (1.00 to 1.00) | 0.479 | 1.00 (1.00 to 1.00) | 0.738 |
| ↓ Stroke work indexed to height<sup>2.7</sup> | 1.00 (1.00 to 1.00) | 0.528 | 1.00 (1.00 to 1.00) | 0.600 |
| ↓ Stroke work indexed to LV mass | 1.01 (1.00 to 1.01) | 0.024 | 1.00 (1.00 to 1.01) | 0.111 |
| ↓ Global longitudinal strain | 1.10 (1.03 to 1.17) | 0.003 | 1.09 (1.02 to 1.16) | 0.013 |
| ↓ GLASE | 1.00 (1.00 to 1.00) | 0.348 | 1.00 (1.00 to 1.00) | 0.413 |
| ↓ GLASE indexed to BSA | 1.00 (1.00 to 1.01) | 0.118 | 1.00 (1.00 to 1.01) | 0.281 |
| ↓ GLASE indexed to height<sup>2.7</sup> | 1.01 (1.00 to 1.02) | 0.151 | 1.01 (1.00 to 1.02) | 0.256 |
| ↓ GLASED | 1.38 (1.11 to 1.73) | 0.005 | 1.28 (1.01 to 1.63) | 0.038 |

